# Spatially constrained ICA enables robust detection of schizophrenia from very short resting-state fMRI data

**DOI:** 10.1101/2022.03.17.22271783

**Authors:** Marlena Duda, Armin Iraji, Judith M. Ford, Kelvin O. Lim, Daniel H. Mathalon, Bryon A. Mueller, Steven G. Potkin, Adrian Preda, Theo G.M. Van Erp, Vince D. Calhoun

## Abstract

Resting-state functional network connectivity (rsFNC) has shown utility for identifying characteristic functional brain patterns in individuals with psychiatric and mood disorders, providing a promising avenue for biomarker development. However, several factors have precluded widespread clinical adoption of rsFNC diagnostics, namely a lack of standardized approaches for capturing comparable and reproducible imaging markers across individuals, as well as the disagreement on the amount of data required to robustly detect intrinsic connectivity networks (ICNs) and diagnostically relevant patterns of rsFNC at the individual subject level. Recently, spatially constrained independent component analysis (scICA) has been proposed as an automated method for extracting ICNs standardized to a chosen network template while still preserving individual variation. Leveraging the novel scICA methodology, which solves the former challenge of standardized neuroimaging markers, we investigate the latter challenge of identifying a minimally sufficient data length for clinical applications of resting-state fMRI (rsfMRI). Using a dataset containing individuals with schizophrenia and controls (*M* = 310) as well as simulated rsfMRI, we evaluate the robustness of ICN and rsFNC estimates at both the subject- and group-level, as well as the performance of diagnostic classification, with respect to the length of the rsfMRI time course. We found individual estimates of ICNs and rsFNC from the full-length (5 minute) reference time course were sufficiently approximated with just 3-3.5 minutes of data (*r* = 0.85, 0.88, respectively), and significant differences in group-average rsFNC could be sufficiently approximated with even less data, just 2 minutes (*r* = 0.86). The results from the shorter clinical data were consistent with the results from the longer simulated data, reliably estimating both individual- and group-level metrics from the full-length (30 minute) reference with just 3-4 minutes of data (*r* = 0.85 - 0.88). Furthermore, we found a model trained on 2 minutes of data retained 97-98% classification accuracy relative to that of the full-length reference model. Our results suggest that clinical rsfMRI scans, when decomposed with scICA, could potentially be shortened to just 2-4 minutes without significant loss of individual rsFNC information or classification performance of longer scan lengths.

## 1. INTRODUCTION

Resting-state functional MRI (rsfMRI) has been a valuable tool for identifying and investigating brain networks and their functional interactions, often referred to as resting-state functional network connectivity (rsFNC), in both typical individuals and those diagnosed with psychiatric and mood disorders. Clinically, rsfMRI offers several benefits, namely that it is non-invasive, it is relatively easy to administer, and imposes fewer demands on patients than other imaging techniques or task-based fMRI paradigms, an important consideration for clinical populations that may not be able to perform standardized tasks in the scanner. Studies of rsFNC have also identified characteristic and reproducible connectivity patterns capable of discriminating between various diagnostic groups [1]–[3], as well as “fingerprinting” individuals and predicting behavior [4].

While these benefits show promise for rsFNC to serve as a potential biomarker and move towards precision diagnosis in the currently tangled landscape of psychiatric disorders, several factors have prevented widespread clinical adoption of such methods. One such challenge is the lack of standardized approaches for capturing imaging markers, in this case individualized intrinsic connectivity networks (ICNs), that are reproducible and comparable across individuals. Independent components analysis (ICA) is a widely used data-driven approach for extracting maximally spatially independent components that share co-varying activation patterns from voxel-level fMRI data, and though several group ICA methods have been developed that enforce correspondence between individual-level ICNs in a given group analysis [5]–[7], there is no such guarantee of correspondence across different datasets. To address this challenge, spatially constrained ICA (scICA) methods have recently been proposed [8] that can extract individualized ICNs guided by the spatial prior of an independently derived and validated network template. The scICA approach is fully automated and ensures the correspondence of ICNs across subjects while maintaining individualized identification of components, suggesting it can be of great use for precision biomarker development.

In addition, there is currently debate in the field surrounding the amount of rsfMRI data needed to generate robust estimates of functional networks and the corresponding resting-state functional connectivity (rsFC) between them. Typically, rsfMRI scans lengths range from 5-15 minutes, but recent work has yielded conflicting results for “optimal” scan lengths, suggesting as little as 5-6 minutes [9]–[11] or as much as 30-40 minutes [12]–[14] of data are necessary to produce sufficiently reliable estimates of individual rsFC. While shorter scanning sessions would be more cost- and resource-efficient for clinical implementation, the ICN and rsFC estimates from shorter time courses can be more susceptible to spurious noise; conversely, while longer scanning sessions have the benefit of averaging across more data, the longer a subject spends in the scanner the more susceptible they are to fatigue, increased head motion, drowsiness, and fluctuations in vigilance [15], which also contribute to noise. Furthermore, individuals with psychiatric or mood disorders can become distressed in the MRI scanner and be unable to tolerate long acquisitions, making the scan duration an increasingly important factor for development of neuroimaging biomarkers with practical clinical utility. Thus, the lack of consensus around an appropriate “minimally sufficient” scan length for clinical applications of rsfMRI has left this an open area of research.

Importantly, existing studies of scan length reliability have focused on atlas- and seed-based approaches. To the best of our knowledge there has been no such examination of reliability using a data-driven ICA approach, specifically scICA. We suggest that the scan length required to achieve reliable results is at least in part dependent on the methodological approach employed. Thus, a data-driven method may produce results that differ from seed- and atlas-based approaches. Moreover, we hypothesize that the regularization provided by the spatial priors in scICA serve to stabilize the independent component solution even when less data is used, providing higher reliability at shorter scan lengths than what has been reported for non-ICA approaches. Motivated by the lack of consensus in recommended scan lengths for clinical applications, we investigate the robustness of both subject-specific ICNs extracted via scICA and their resultant rsFNC matrices with respect to time series length. As we are interested specifically in studying minimal sufficient scan lengths in the context of clinical biomarker development, we also evaluate the robustness of scan length to the identification of significant group differences in rsFNC between, as well as classification of, schizophrenia and control groups. To supplement the relatively short clinical scans and study the effect of reference length on minimal sufficient scan lengths, we replicate these experiments using simulated 30-minute rsfMRI time courses as the reference.

## 2. MATERIALS AND METHODS

### 2.1 Spatially Constrained ICA

A spatially constrained ICA (scICA) approach called multivariate-objective optimization ICA with reference (MOO-ICAR) was implemented using the GIFT software toolbox (http://trendscenter.org/software/gift) [16]. Briefly, the MOO-ICAR framework estimates subject-level independent components (ICs) using existing network templates as spatial guides [8]. In this work, we utilized the NeuroMark template (described in [8] and available at https://trendscenter.org/data/) (Figure 1), which consists of *N* = 53 ICNs categorized into seven functional domains: subcortical (SC), auditory (AUD), sensorimotor (SM), visual (VIS), cognitive-control (CC), default mode (DM) and cerebellar (CB). The following equation represents how the l^th^ network can be estimated for the k^th^ subject using the network template *S*_*l*_ as guidance:

**Figure 1.**
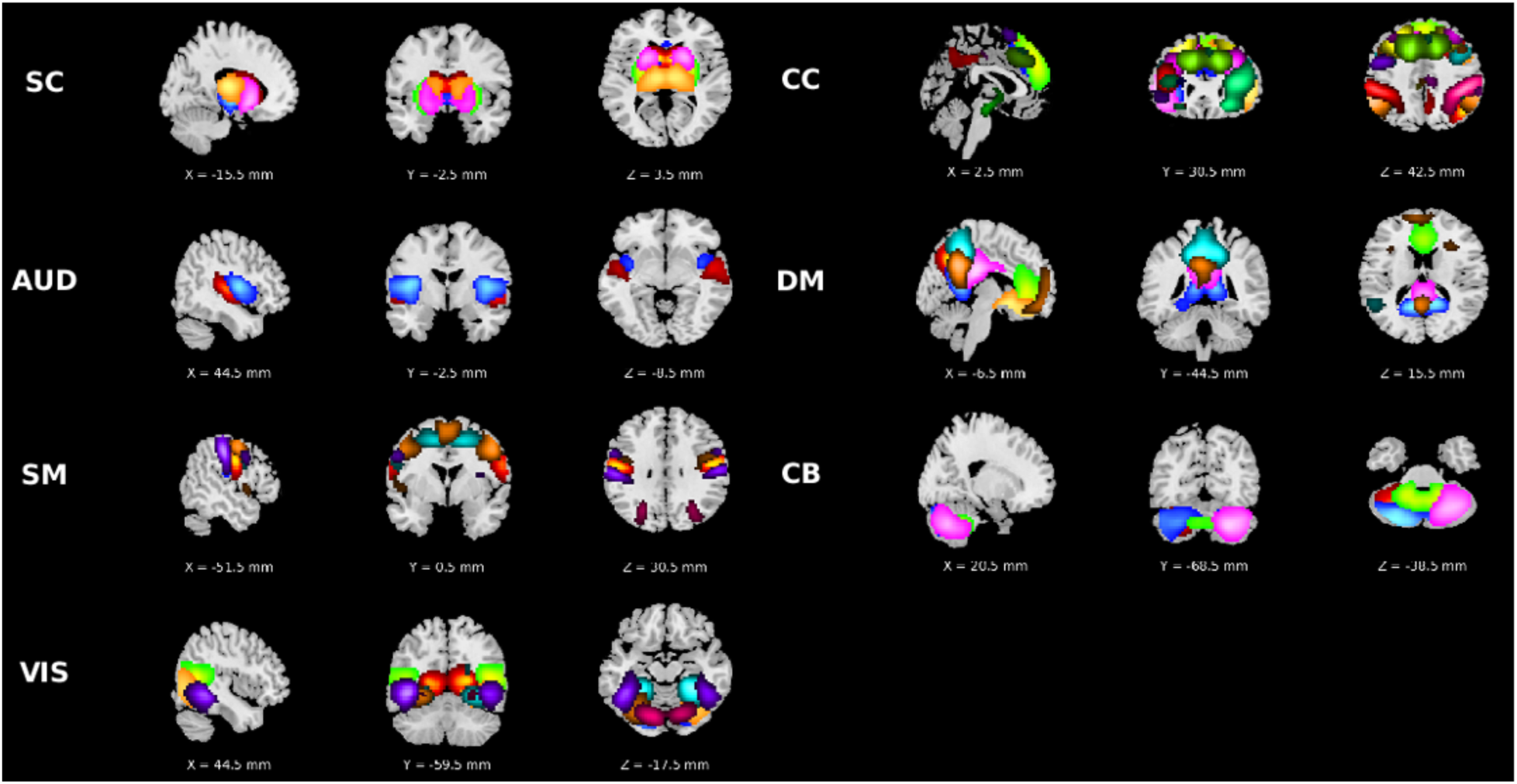
NeuroMark network template used for spatially constrained ICA. Domain abbreviations: subcortical (SC), auditory (AUD), sensorimotor (SM), visual (VIS), cognitive-control (CC), default mode (DM) and cerebellar (CB).

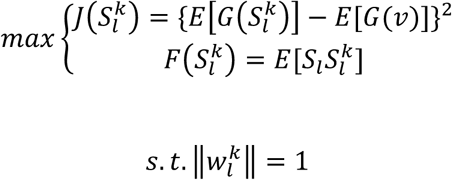

In this formulation, 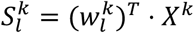 represents the estimated l^th^ network of the k^th^ subject, where *X*^*k*^ is the whitened fMRI data matrix of the kth subject and 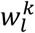 is the unmixing column vector, to be solved in the optimization functions. The function 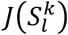 serves to optimize the independence of 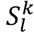 via negentropy. Here, v is a Gaussian variable with mean zero and unit variance, G() is a nonquadratic function, and E[] denotes the expectation of the variable. The function 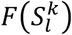 serves to optimize the correspondence between the template network (*S*_*l*_) and subject network 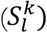. The optimization problem is solved by applying a linear weighted sum to combine the two objective functions, with weights set at 0.5. Applying scICA via MOO-ICAR to each scan extracts subject-specific ICNs corresponding to each of the *N* network templates, along with the relevant time courses.

### 2.2 Evaluation Framework

A flowchart of our analysis framework is presented in Figure 2. We began by partitioning the preprocessed rsfMRI data into incrementally longer segments, beginning with the first 1 minute, then 2 minutes, and so on until the full length of the data was reached. Next, we applied scICA via the MOO-ICAR framework (Section 2.1) separately to each length of rsfMRI, extracting subject-specific ICNs and their corresponding TCs. Results from the full-length TC were considered the reference or “gold standard” in our evaluation, thus the robustness of shorter data lengths was evaluated with respect to the full TC. To evaluate the spatial map stability of the scICA decomposition, as well as the rsFNC stability of their respective TCs, we computed subject-level Pearson correlations between full TC and partial TC experiments for both spatial composition of the extracted ICNs and the resultant rsFNC matrices. We utilized a robustness threshold of correlation ≥ 0.85 to the reference (i.e., full-length TC), formerly proposed in [12], to identify a “minimally sufficient” data length with respect to these metrics. Pearson’s r was used to assess reliability rather than intraclass correlation (ICC) to enable direct comparison to, and use of the same evaluation criteria of, this prior work.

**Figure 2.**
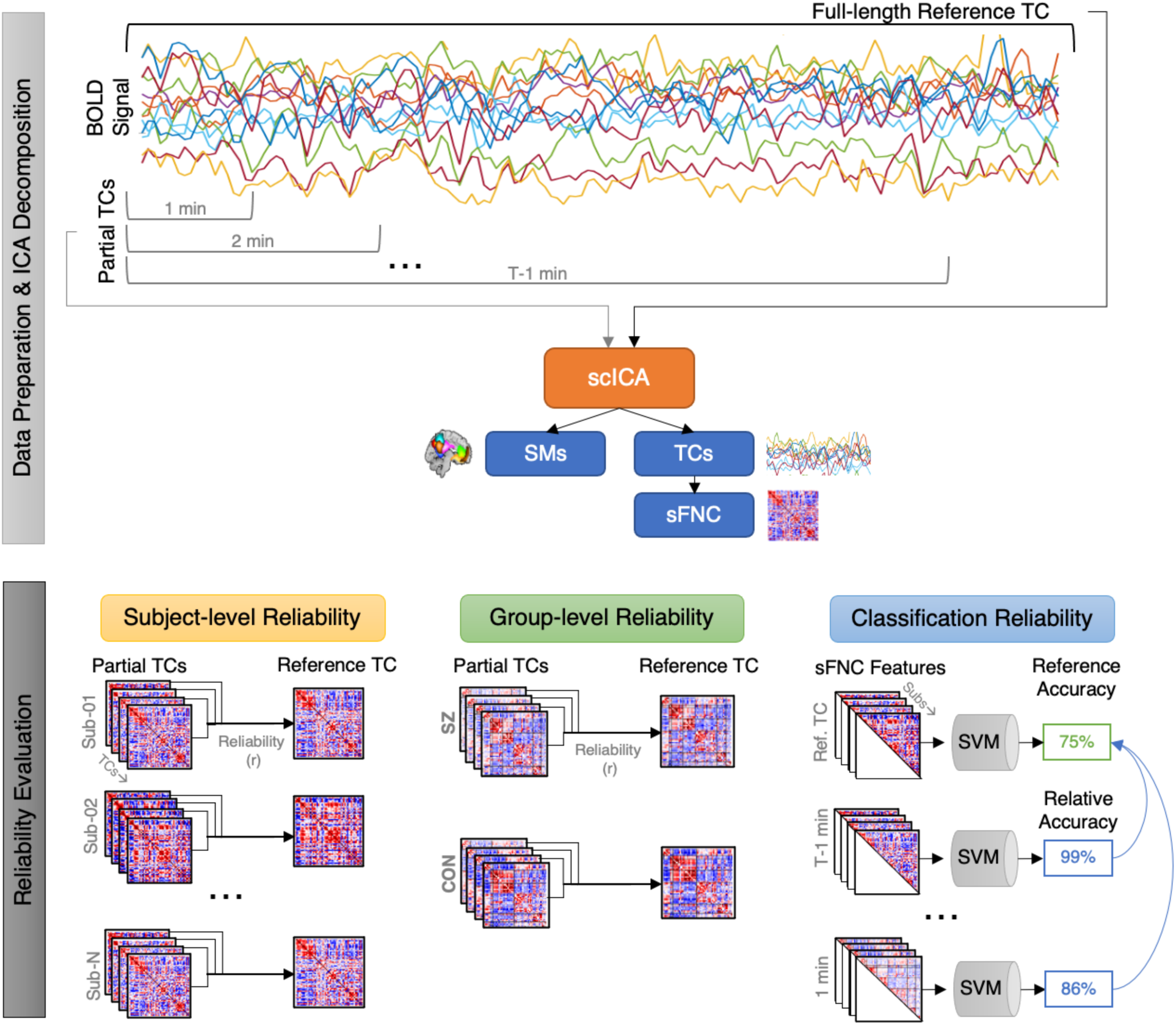
Analysis framework. Data preparation steps including extraction of partial time courses, as well as spatially constrained ICA and its outputs are depicted on the top panel. Reliability evaluation steps including subject-level, group-level and classification reliability experiments are depicted on the bottom panel. Abbreviations: time course (TC); blood oxygen level-dependent (BOLD); independent components analysis (ICA); spatially constrained ICA (scICA); spatial maps (SMs); static functional network connectivity (sFNC); schizophrenia (SZ); control (CON); support vector machine (SVM).

In each experiment, subject-level static functional network connectivity (sFNC) was computed via pairwise Pearson correlation between time courses of all ICNs, resulting in an *N* × *N* sFNC matrix. Fisher’s Z transform was applied to all sFNC matrices to improve normality. We computed group-average sFNC matrices for the schizophrenia (SZ) and control (CON) groups (or groups A and B in the simulated data) at each data length. We investigated group differences in sFNC values between the SZ and CON (or Group A/B) populations using two parallel approaches—two-sample t-test as well as univariate multiple linear regression including age, gender, scanning site and head motion as covariates. Significant group differences in sFNC were identified as relationships whose p-values survive FDR correction at α_FDR_ = 0.05. To further evaluate the stability of diagnostically relevant rsFNC patterns as the length of fMRI data decreased, we computed group-level correlations between the full TC and each of the partial TCs for each of the mean SZ/CON sFNC matrices, t-test t-values, and diagnosis term t-values from the multiple linear regressions.

We apply this evaluation framework to both our discovery rsfMRI dataset (*M* = 310 subjects) as well as our simulated dataset (*M* = 100 subjects; Section 2.4).

### 2.3 Group Classification

We further investigate the robustness and clinical utility of scICA-based estimates of rsFNC with a group classification task. Using our discovery rsfMRI dataset as training data (*M* = 310 subjects; 150 SZ), we generated subject-level feature vectors for each data length by extracting the upper triangular of the corresponding scICA-derived sFNC matrix. Using the “fitclinear” function in MATLAB, we fit binary LASSO-regularized linear SVM classification models separately for each data length (1, 2, 3, 4, and 5 minutes) to classify each subject as SZ/CON. For each model, the lambda parameter was tuned using five-fold cross validation. After obtaining the optimal lambda value, performance for each of the five models was estimated with 500 rounds of bootstrap resampled five-fold cross validation. The final five models were trained on the full training dataset and tested on the held-out independent validation dataset (*M* = 129 subjects; 50 SZ) using the same bootstrap resampling scheme (500 rounds) for external evaluation of classification performance and generalizability at each data length.

### 2.4 Data and Preprocessing

#### 2.4.1. Clinical fMRI Data

We utilized an age- and gender-matched dataset including 150 individuals with schizophrenia (SZ) and 160 controls (CON) [17]. Resting-state fMRI (rsfMRI) data were collected with 3-Tesla MRI scanners with a repetition time (TR) of 2 seconds, voxel size of 3.44 × 3.44 × 4 mm, a slice gap of 1 mm, and a total of 157 volumes. Subjects were instructed to keep their eyes closed during the resting-state scan but not fall asleep. Additionally, we utilized an independent validation dataset in our classification experiments, consisting of 50 SZ and 79 CON samples [18]. The rsfMRI data were collected with 3-Tesla MRI scanners with a TR of 2 seconds, voxel size of 3.75 × 3.75 × 4.55 mm, and a total of 145 volumes. In the validation set, subjects were instructed to keep their eyes open and passively stare at a fixation cross. Informed consent was obtained from each participant prior to scanning and all studies were approved by the Institutional Review Boards of the corresponding institutions (University of California: Irvine, San Diego, Los Angeles; Stanford University, University of New Mexico, University of Iowa, University of Minnesota, Duke University, University of North Carolina, Brigham and Women’s Hospital, Massachusetts General Hospital, Yale University). For both data sets preprocessing included brain extraction, slice-timing, and motion correction steps. Preprocessed data were then registered into structural MNI space, resampled to 3 mm^3^ isotropic voxels, and spatially smoothed using a Gaussian kernel with a 6 mm full-width at half-maximum (FWHM) on a per-subject basis. Finally, the first five timepoints were trimmed from the time course and all voxel time courses were z-scored.

#### 2.4.2 Simulated Data

To supplement the relatively short data lengths available from our clinical rsfMRI datasets, we simulated a set of longer fMRI time courses using the SimTB toolbox [19], (freely available for download at https://trendscenter.org/software/simtb/). Briefly, SimTB utilizes a data generation model under the assumption of spatiotemporal separability, meaning the simulated fMRI data can be expressed as the product of time courses (TCs) and spatial maps (SMs). These simulated data are modeled with realistic dimensions, spatiotemporal activations, and noise characteristics typical of fMRI datasets [20]. For subjects *i* = 1, …, *M*, we model *C* components, each consisting of a SM and corresponding TC. In our simulation, we set *M* = 100 subjects and *C* = 29 components. SMs have *V* = 148 × 148 voxels and TCs are *T* = 900 time points in length with a TR = 2 seconds (totaling 30 minutes of data). The *C* = 29 components used in our simulation are shown in Figure 3. In our simulation, individual variability in component SMs is implemented as follows. SMs are translated according to a bivariate normal distribution with mean zero and standard deviation of 0.75 voxels and are rotated by normally distributed angles with mean zero and a standard deviation of 1º. SM size is also randomly varied, with the spread parameter, *ρ*, uniformly distributed between 0.85 and 1.15. Additionally, component amplitudes, *g*_*ic*_, are distributed normally with a mean of 3 and standard deviation of 0.3, simulating individual TC variation.

**Figure 3.**
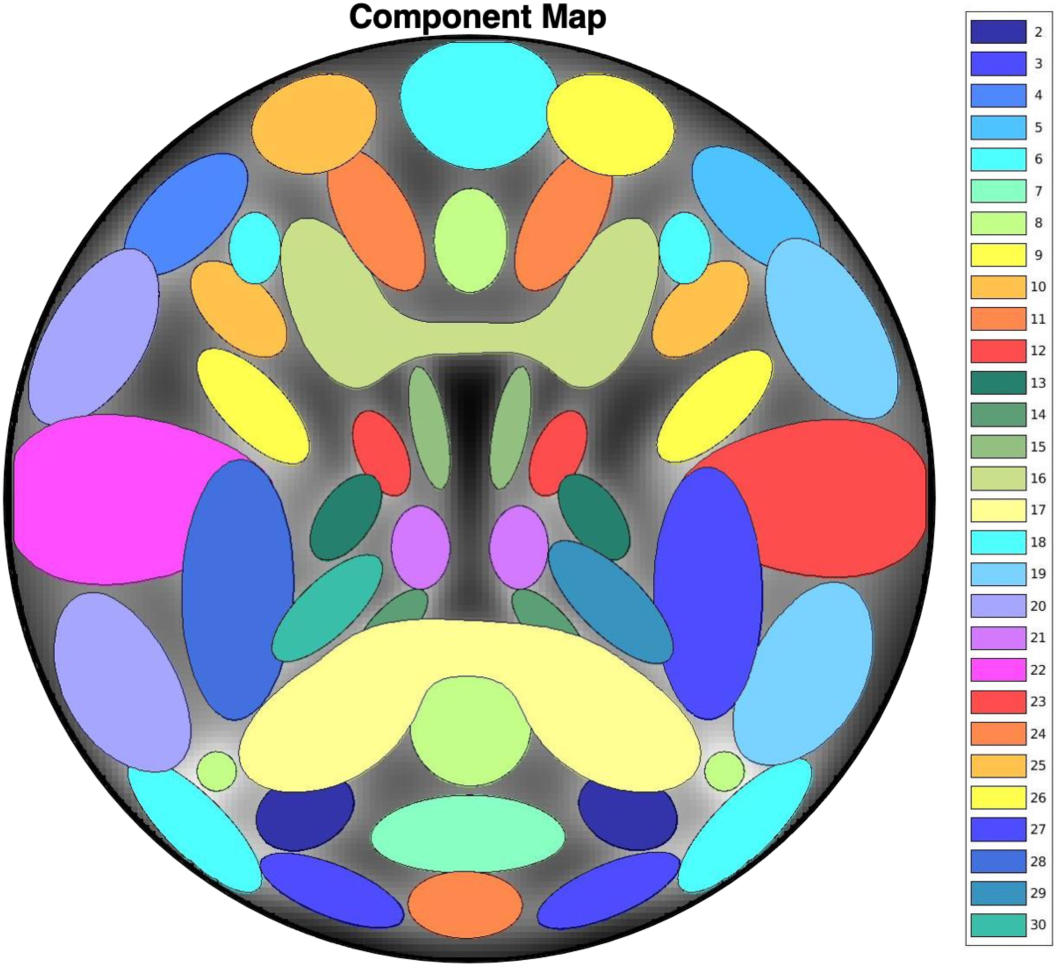
Spatial map of the 29 components used in our simulation study (corresponding to default components numbers 2-30 in the SimTB software).

We model two groups, A and B, that differ in four ways, modeled after a similar experiment in [20] and described in detail below. Each group has 50 subjects, for *M* = 100 subjects total. Group differences are as follows. (1) Group A has larger amplitude for component 7 than Group B. This is modeled by distributing *g*_*i7*_ as normal with mean 3.5 and standard deviation 0.3 for Group A, and mean 2.5, standard deviation 0.3 for Group B. (2) Groups A and B have different shapes for a network composed of components 5 and 10. The network is modeled by assigning shared events between components 5 and 10 and setting the amplitude of unique events, *A*_*u*_ = 0, creating identical TCs. For Group A, the amplitude of component 5 is *g*_*i5*_ = 0.7 × *g*_*i10*_, whereas for Group B *g*_*i5*_ = 1 × *g*_*i10*_. Thus, Group A has a network where the component 5 node is weaker than the component 10 node and Group B has two nodes of equivalent strength. (3) Groups A and B have different shapes and different amplitudes for a network composed of components 22 and 23. For Group A, *g*_*i23*_ = 0.7 × *g*_*i22*_, where *g*_*i22*_ is distributed normally with mean 6 and standard deviation 0.3. For Group B, *g*_*i23*_ = 1 × *g*_*i22*_, where *g*_*i22*_ is distributed normally with mean 3 and standard deviation 0.3. Thus, Group A has a lateralized network where the left node is stronger than the right and Group B has a bilaterally symmetric network. Furthermore, the amplitude of the network for Group A is much larger than the amplitude for Group B. (4) Group B has stronger FNC between components 3 and 4 than Group A. This is modeled by first designating shared events between components 3 and 4, then distributing *A*_*u*_ as uniform between [0.5,1.0] for Group A and between [0.65, 1.15] for Group B.

## 3. RESULTS

### 3.1 Subject-level estimates of ICNs and sFNC are highly robust to data length

We evaluated the stability of subject-level estimates of ICN spatial maps and sFNC matrices derived via scICA with respect to the length of rsfMRI data (Figure 4A-B). We observed increasing variation in the stability of subject-level sFNC estimates at shorter data lengths, but comparatively little variation in the ICN spatial stability, likely owing to the regularization provided by the spatial priors in the scICA decomposition. Similarly, we found no discernible group differences between SZ and CON in ICN spatial map stability, whereas the CON group exhibited slightly higher stability in sFNC than the SZ group at shorter data lengths. Results showed only 3 minutes of rsfMRI data were sufficient to meet the robustness threshold for the subject-specific ICN spatial maps and 3.5 minutes were sufficient for the corresponding subject-level sFNC estimates.

**Figure 4.**
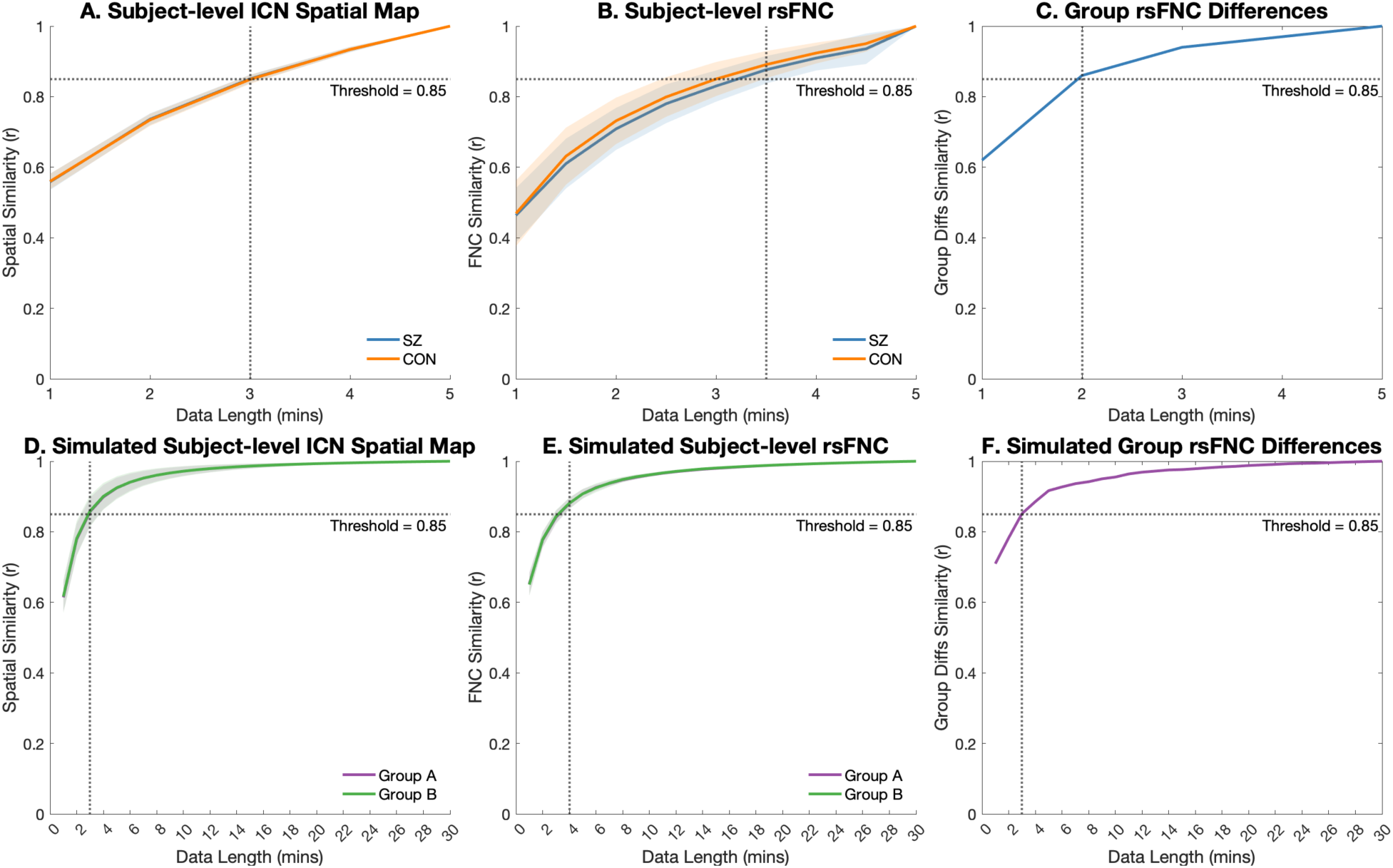
Reliability results for clinical (A-C) and simulated (D-F) datasets. Subject-level measures (A-B, D-E) show mean (solid line) and standard deviation (shaded area) across all subjects. Dotted lines indicate data lengths at which the measures meet or exceed the robustness threshold (r ≥ 0.85 [12]). Abbreviations: intrinsic connectivity network (ICN); resting-state functional network connectivity (rsFNC); schizophrenia (SZ); control (CON).

### 3.2 Group-level sFNC patterns can be reliably estimated with less data

In addition to subject-level reliability of ICN and sFNC estimates, we evaluated the reliability of group-level sFNC patterns across TC lengths. Results showed the characteristic rsFNC signatures for the SZ and CON groups were highly robust to the length of rsfMRI data used (Figure 5A-B), indicating high group-level rsFNC stability. The group-level sFNC matrices derived from even 1 minute of data were highly correlated to the full TC reference for both the SZ (r = 0.94) and CON (r = 0.93) groups, and this relationship continually increased as more of the rsfMRI time course was utilized.

**Figure 5.**
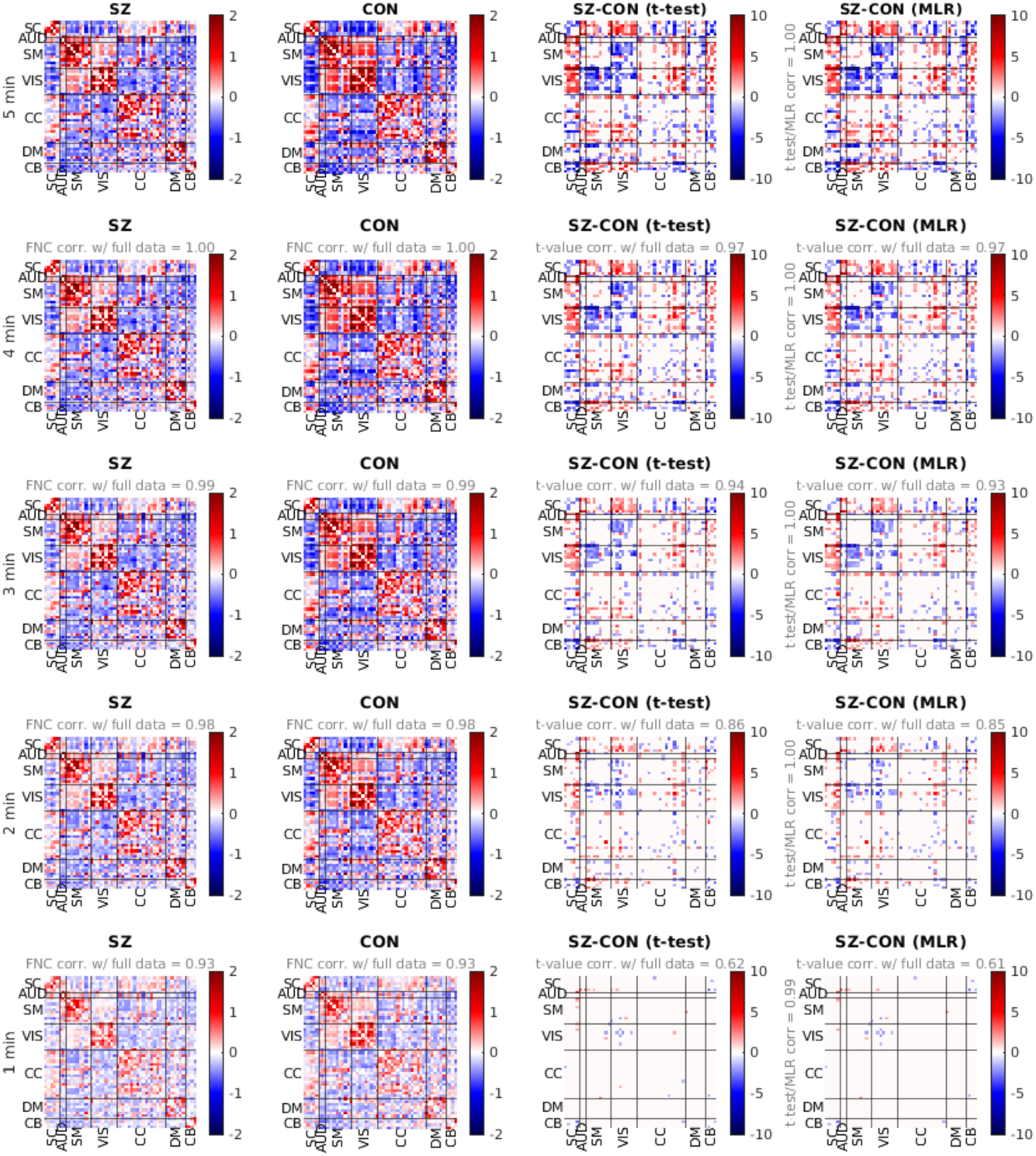
(A-B) Mean functional network connectivity (FNC) for patients with schizophrenia (SZ) (A) and typical controls (CON) (B) from the full (5 min) and partial (1 – 4 min) fMRI time course. For experiments on partial time series data, Pearson correlation with FNC from the full data is reported. (C-D) Group differences (SZ-CON) in FNC. Values are plotted as −log10(p-value) × sign(t-value), where statistics are obtained via t-test across diagnosis groups (C) or from the diagnosis term in univariate multiple regression (MLR) models (D). For experiments using partial time series data, t-value correlation with full data experiments is reported.

We found significant group differences in sFNC patterns across all experiments, both with the classic t-test and with multiple regression with added covariates (Figure 5C-D). Results showed near-perfect concordance between these two parallel methodologies, with correlations in t-values ≥ 0.99 in each experiment. We found only 2 minutes of rsfMRI data were required to meet the robustness threshold (correlation ≥ 0.85 to the reference) and reliably estimate the significant group differences identified from the full TC (Figure 4C). Specifically, the results showed that the group differences that were most robust to data length were lower within-domain connectivity of the VIS domain and higher cross-domain connectivity between SC-SM and SC-VIS domains in the SZ group compared to that of the CON group.

### 3.3 Simulation study suggests ICN and sFNC estimates from relatively long rsfMRI can be reliably estimated from very short TCs

To evaluate how reliability is affected with longer rsfMRI as the reference, we simulated 30 minutes of rsfMRI data designed to model two groups with distinct activation and FNC patterns (Section 2.4.2). The stability of subject-level estimates of ICN spatial maps and sFNC matrices derived via scICA with respect to the length of the simulated rsfMRI data is shown in Figure 4D-E. Somewhat contrary to results of the SZ/CON data, we observed slightly larger variation in subject-level spatial map stability at shorter data lengths compared to that of subject-level sFNC. This could be due in part to the induced variability in translation/rotation/size of the simulated ICN spatial maps. The results showed that the robustness threshold of mean correlation ≥ 0.85 to the reference was reached with 3 minutes of data for the spatial composition of subject-specific ICNs and 4 minutes of data for subject-level sFNC, corresponding well to the results from the clinical data and suggesting individualized rsFNC features averaged across even relatively long TCs can be reliably estimated from just a fraction of the time series. Group-level mean sFNC was highly robust to data length, and similarly we found 3 minutes of data were required to reach the robustness threshold for significant group differences in sFNC (Figure 4F; Figure 6).

**Figure 6.**
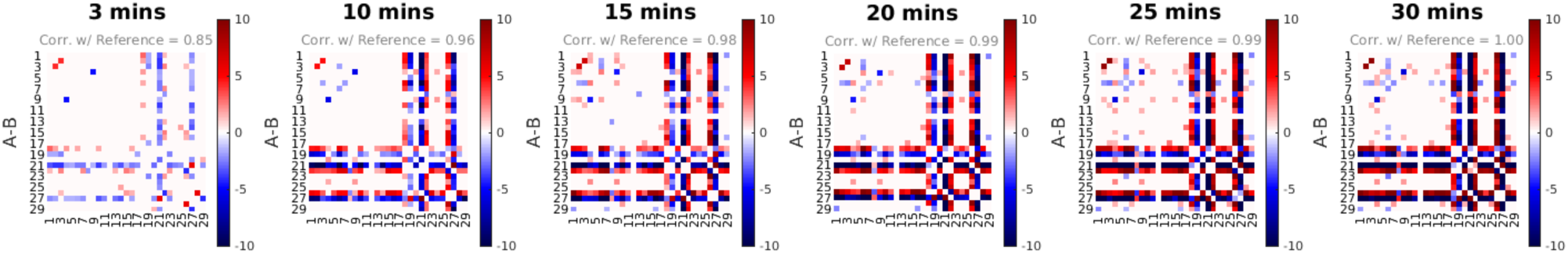
Group differences in rsFNC between Groups A and B in the simulated dataset across various data lengths. Values are plotted as −log10(p-value) × sign(t-value).

### 3.4 Classification accuracy remained stable as less rsfMRI data used for prediction

The results from our classification experiments showed highly stable classification accuracy for models fit across the range of 2-5 minutes, with the model trained on 2 minutes of data attaining a relative performance to that of the full TC reference model of 98% in the internal cross validation (0.73 vs 0.74) and 97% in the external validation (0.68 vs 0.70) (Figure 7A). A full report of mean classification performance metrics including accuracy, AUC, sensitivity and specificity is listed in Table 1. Notably, the results showed that 4 minutes of data had a relative performance of 97%-100% compared to the reference TC across all four classification metrics, followed by 95-100% for 3 minutes of data, 94-100% for 2 minutes of data, and 79-90% for just 1 minute of data. These results suggest that in addition to being robust on an individual level, sFNC features derived from scICA decomposition of rsfMRI data capture clinically relevant patterns of rsFNC, even from very short scan lengths, and yield diagnostic classifications that are highly robust to data length.

**Table 1.** Classification results. Report of the mean classification accuracy, area under the receiver operating characteristic curve (AUC), sensitivity and specificity for both internal cross validation (CV) and external testing experiments for SVM models trained on features derived from all data lengths (1 – 5 minutes). We report both the observed (Obs.) values as well as the relative (Rel.) values compared to that of the full-length reference time course (5 minutes).

|  | Internal 5-fold CV Results (500x Bootstrap) |  |  |  |  |  |  |  | External Validation Results (500x Bootstrap) |  |  |  |  |  |  |  |
| --- | --- | --- | --- | --- | --- | --- | --- | --- | --- | --- | --- | --- | --- | --- | --- | --- |
|  | Accuracy |  | AUC |  | Sensitivity |  | Specificity |  | Accuracy |  | AUC |  | Sensitivity |  | Specificity |  |
|  | Obs. | Rel. | Obs. | Rel. | Obs. | Rel. | Obs. | Rel. | Obs. | Rel. | Obs. | Rel. | Obs. | Rel. | Obs. | Rel. |
| 5 min. | 0.74 | - | 0.83 | - | 0.71 | - | 0.78 | - | 0.70 | - | 0.78 | - | 0.70 | - | 0.70 | - |
| 4 min. | 0.73 | 0.99 | 0.82 | 0.98 | 0.69 | 0.97 | 0.78 | 1.00 | 0.70 | 0.99 | 0.78 | 1.00 | 0.69 | 0.99 | 0.71 | 1.00 |
| 3 min. | 0.74 | 1.00 | 0.81 | 0.97 | 0.70 | 0.99 | 0.78 | 1.00 | 0.68 | 0.98 | 0.76 | 0.97 | 0.67 | 0.95 | 0.71 | 1.00 |
| 2 min. | 0.73 | 0.98 | 0.80 | 0.97 | 0.67 | 0.95 | 0.78 | 1.00 | 0.68 | 0.97 | 0.74 | 0.94 | 0.67 | 0.96 | 0.68 | 0.97 |
| 1 min. | 0.63 | 0.85 | 0.69 | 0.83 | 0.56 | 0.80 | 0.70 | 0.90 | 0.57 | 0.82 | 0.65 | 0.82 | 0.56 | 0.79 | 0.60 | 0.85 |

**Figure 7.**
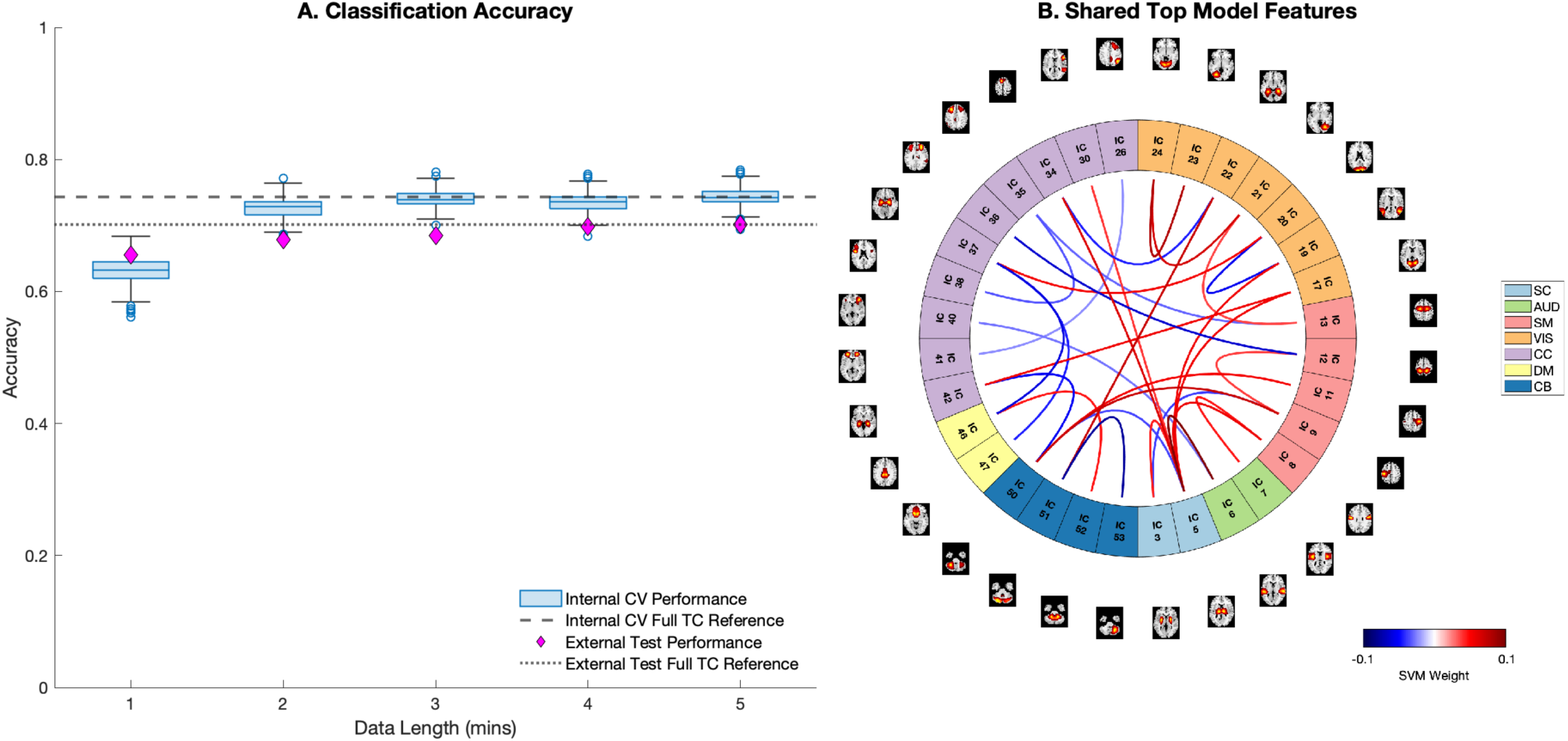
(A) Classification performance of linear SVM models trained at each data length. (B) FNC edges in the top 20% of SVM feature weights shared among all five models.

For each of the five models we also extracted the most influential sFNC edges, defined as the top 20% of features with the highest magnitude weights. We found 31 sFNC edges were commonly included among the top feature weights across all five models (Figure 7B), which mainly belonged to the subcortical (SC), visual (VIS) and sensorimotor (SM) domains. These results align with the significant results of our group differences analysis and involve domains that have previously been implicated in relation to schizophrenia in the literature [21], [22].

## 4. DISCUSSION

There is increasing agreement within the psychiatric field that current behavioral diagnostic criteria are likely insufficient to properly disentangle the complex and blurred boundaries between psychiatric disorders as they are presently defined. In light of this, much research is focused on the identification and development of biomarkers for psychiatric disorders, with considerable attention on neuroimaging-based methods, such as rsfMRI [3]. Resting-state fMRI approaches are well suited for clinical biomarkers, not only due to their non-invasive nature and relatively high spatial and temporal resolution, but also due to the low demand placed on patients during resting-state acquisitions, which is especially important for use in psychiatric populations that may experience difficulty performing structured cognitive tasks in the MRI scanner. Another key consideration in the development of psychiatric rsfMRI biomarkers is the duration of image acquisition required. Minimizing the time in the scanner is critical, both for maximizing the comfort of patients and for resource efficiency of high-demand MRI equipment. However, lack of consensus for a minimally sufficient rsfMRI scan length has factored into the delayed adoption of neuroimaging biomarkers in clinical settings. Previous reports have produced conflicting results, proposing that as little as 5-6 minutes [9], [10] and as much as 30-40 minutes [12]– [14] are necessary to produce robust and reliable estimates of individual rsFNC, with most studies most recent works advocating for longer acquisitions [23].

Here, we contribute to this debate by providing an evaluation of reliability of both subject- and group-level measures of rsFNC for individuals with schizophrenia and controls with respect to the length of rsfMRI data used for analysis. Our work differs from existing studies mainly in our use of a spatially constrained ICA approach, which leverages existing network templates as spatial priors to guide the spatiotemporal decomposition of the fMRI data. The data-driven nature of scICA enables individualized identification of ICNs while simultaneously ensuring their correspondence to a validated set of functional networks of interest and automatedly discarding noise components. We hypothesized that the additional regularization provided by scICA may serve to enable robust and reliable estimation of rsFNC at shorter scan lengths than those previously reported in when utilizing atlas- or seed-based analyses. Beyond regularization, the scICA has several additional benefits that are favorable for use in clinical settings. Firstly, scICA is completely automated, negating the need for any manual annotation of relevant brain regions from noise components. Secondly, scICA provides crucial correspondence in components between subjects, allowing it to directly patch into any downstream steps in the biomarker algorithm without the need for intermediary steps such as computation of spatial overlaps. Thirdly, scICA is fully parallelizable and can be applied to each patient’s scans independently without the need for group analysis. Lastly, scICA can be used with any customized spatial templates that contain ICNs of interest, allowing for additional tailoring to the relevant networks for any given diagnosis.

Overall, our results showed that both subject-level and group-level sFNC features could be reliably estimated from a fraction of the full length rsfMRI time series. Specifically, we found that just 3-4 minutes of data sufficiently surpassed the chosen robustness threshold for approximating individual rsFNC and ICNs from the reference TC. Importantly, this same threshold was used for evaluation in a previously published seed-based study of scan length reliability, but yielded a much different result, reporting that at least 30 minutes of rsfMRI data were required for the threshold to be reliably met [12]. In our study, we found the minimum sufficient scan length in the 3-4 minute range held true when the reference sFNC estimates were derived from both shorter (5 minutes) or longer (30 minutes) TCs alike, suggesting the conflicting results between the current and previous reports cannot be attributed to longer reference lengths alone.

Our study also differs from existing work in this area because it goes beyond individual-level reliability and evaluates robustness of group-level metrics as well as diagnostic classification performance, directly evaluating scan length reliability in the context of clinical utility. We found that even less data was required to reliably approximate significant group differences in rsFNC than at the subject level (2 minutes vs. 3.5 minutes in clinical experiments, 3 vs. 4 minutes in simulated experiments), which is expected due to the benefit of group averaging. Surprisingly, we also found high reliability in classification performance at very short lengths, achieving a relative diagnostic accuracy of 97% compared to the full-length reference TC with just 2 minutes of data. This result was somewhat unexpected, as classification of individuals is a much harder task than identifying group differences in aggregate. However, we found that even if overall rsFNC robustness was not reached with just 2 minutes of data, the rsFNC features that are generated at this very short scan length do indeed capture clinically relevant patterns of subject-level rsFNC and yield diagnostic classifications that closely resemble those of the full reference TC. While these results highlight important differences in rsFNC between individuals with schizophrenia and controls that can be identified even in very short fMRI scans, the results should be interpreted with caution given the history of medication in the schizophrenia group.

This study is limited by the acquisition protocols of the clinical data sets used in the analyses, which consist of relatively short (∼5 minutes) one-time scans of individuals. Longer scans would provide more flexibility to test a wider range of data lengths in our reliability analysis, increasing confidence in the results. Furthermore, longitudinal data consisting of multiple scanning time-points would enable a higher-level assessment of reliability across days or weeks, accounting for intra-individual variation that may occur across longer timescales. Future work should assess reliability in other datasets that satisfy these criteria (i.e. the Human Connectome Project (HCP) [24] or Midnight Scan Club (MSC) [12] datasets), to supplement the findings of the present study, which focused on clinically relevant reliability in the context of neuroimaging biomarkers. Furthermore, the results of the simulation study are expectedly limited in 1) the spatial dimensionality, simulating a single axial slice rather than a three-dimensional full brain, 2) strength of engineered group differences, and 3) ability to accurately simulate the properties of true fMRI data. However, the efficacy of the SimTB software has been extensively studied and has shown utility for reliably modeling multi-subject fMRI datasets [19], [20]. Additionally, we only consider the results of scICA with a single network template generated using only control subjects [8]. While most existing reliability studies consider only one brain atlas, a more thorough examination of scICA in the context of reliability and data reduction in the future could include various templates, such as customized templates for certain diagnostic groups. Finally, we acknowledge that the identification of a minimum scan length is dependent upon the metrics of interest, or the evaluation framework used. For example, as our work produced differing results for the evaluation of group-level vs. subject-level rsFNC reliability, future studies that examine other metrics, such as graph theoretic measures of rsFNC (i.e., modularity, global efficiency, etc.), could result in minimum scan recommendations that differ from those in this study.

Our results support the idea that when scICA is employed, a minimally sufficient scan length may exist in the 2-4 minute range that could be favorable for use in clinical settings, both in maximizing clinical efficiency and patient comfort while retaining diagnostic efficacy. However, more work is still required to validate these results in larger (and longer) data sets, as well as in other diagnoses for which rsFNC biomarkers may be useful. Future work may also focus on the benefits of using scICA in the study of time-resolved, or “dynamic” FNC (dFNC). The added regularization afforded by the spatial priors in scICA may help stabilize sliding window estimates of dFNC, potentially increasing the resolution at which dFNC can be studied with the use of smaller window sizes and increasing the reliability of dFNC results overall.

## 5. CONCLUSION

Due to the lack of consensus for minimum fMRI scan length recommendations, specifically for clinical biomarker applications, we studied the robustness of ICN and rsFNC measures with respect to scan length. We found just a fraction (2-4 minutes) of the full-length time series was necessary to sufficiently approximate ICNs and rsFNC at both the subject- and group-level. These findings were consistent across experiments in both shorter clinical data and longer simulated data. Overall, our results suggest clinical rsfMRI scans, when decomposed with scICA, could be shortened to just 2-4 minutes without significant loss of rsFNC information or classification performance of longer scan lengths.

## Data Availability

Details on the availability of the FBIRN dataset used as our discovery dataset can be found at https://www.nitrc.org/projects/fbirn/.
Details on the availability of the COBRE dataset used as our validation dataset can be found at http://fcon_1000.projects.nitrc.org/indi/retro/cobre.html.

## 6. ACKNOWLEDGEMENTS

This work was supported by the National Institutes of Mental Health grant R01MH123610 and National Science Foundation grant 2112455.

